# A Randomized Clinical Trial of Bioimpedance Spectroscopy or Tape Measure Triggered Compression Intervention in Chronic Breast Cancer Lymphedema Prevention

**DOI:** 10.1101/2021.10.12.21264773

**Authors:** Sheila H. Ridner, Mary S. Dietrich, John Boyages, Louise Koelmeyer, Elisabeth Elder, T. Michael Hughes, James French, Nicholas Ngui, Jeremy Hsu, Vandana G. Abramson, Andrew Moore, Chirag Shah

## Abstract

**Background:** This study compared rates of progression to chronic breast cancer-related lymphedema (defined as a ≥ 10% arm volume change from baseline requiring complex decongestive physiotherapy (CDP)) following an intervention for subclinical lymphedema (S-BCRL) triggered by bioimpedance spectroscopy (BIS) or by tape measurement (TM).

**Methods and Results:** This stratified, randomized, international trial enrolled new breast cancer patients undergoing: mastectomy/partial mastectomy, axillary treatment (dissection, sentinel lymph node biopsy >6 nodes or radiation), radiation therapy (chest wall/ breast, supraclavicular fossa), or taxane-based chemotherapy. Following post-surgery eligibility reassessment, centralized, 1:1 randomization to prospective surveillance by BIS or TM occurred. S-BCRL detection triggered a 4-week, 12-hour per day, compression sleeve and gauntlet intervention. The primary outcome (n=209), rates of post-intervention progression to CDP, were assessed over three years. Between June 24, 2014 and September 11, 2018, 1,200 patients were enrolled, 963 randomized (BIS n=482;TM n=481) and 879 analyzed (BIS n=442;TM n=437). Median follow-up was 32.9 months (IQR=22,35). BIS patients triggered an intervention at a lower rate than TM patients (20.1%, n=89 vs 27.5%, n=120, *p* = 0.011). Median months to trigger was longer with BIS than TM (9.7; 95%CI,8.2-12.6 vs 3.9; 95%CI,2.8-4.5, *p* = 0.001). Overall, 14.4%(n=30) progressed post-intervention, with reduced likelihood for BIS patients than TM patients (7.9%, n=7 vs 19.2%, n=23; RR=0.41; 95%CI,0.13-0.81; absolute reduction 11.3%; 95%CI,2.3%-20.3%; *p* = 0.016).

**Conclusions:** As compared to TM, BIS provides a more precise identification of patients likely to benefit from an early compression intervention.

**Condensed Abstract:** This stratified, multi-site, international trial enrolled newly diagnosed breast cancer patients, randomized them to prospective surveillance by bioimpedance spectroscopy (BIS) or tape measurement (TM), and screened them for lymphedema development at frequent intervals for three years after surgery. When subclinical lymphedema was detected a 4-week, 12-hour per day, compression sleeve and gauntlet intervention was implemented. Overall, 14.4% (n=30) progressed post-intervention to chronic lymphedema, with reduced likelihood for BIS patients than TM patients (7.9%, n=7 vs 19.2%, n=23; RR=0.41; 95%-CI,0.13-0.81; absolute reduction 11.3%; 95%-CI,2.3%-20.3%; *p* = 0.016). BIS best supported intervention success for prevention of chronic lymphedema compared to TM.

## Introduction

As breast cancer survivorship rises, the impact of long-term treatment complications have taken on greater significance.^1^ One such complication is breast cancer-related lymphedema (BCRL).^2–4^ Chronic BCRL(C-BCRL) can lead to pain, infections, limited arm function, reduced quality of life, and costly and resource intensive therapies, such as complex decongestive physiotherapy (CDP).^5,6^ Rates of C-BCRL range from 5% with breast conserving surgery with sentinel lymph node biopsy (SNLB) alone to greater than 50% with axillary lymph node dissection (ALND), regional nodal irradiation (RNI), and/or taxane-based chemotherapy.^5^

Identifying patients in the early stage of BCRL can be challenging. Currently, a diagnosis of C-BCRL is usually made after visible/clinically apparent changes or symptoms occur.^7–9^ Prior to these changes, subclinical disease occurs as indicated by an increase in extracellular fluid.^10^ Accurately identifying subclinical disease before it is visible facilitates early intervention and possibly C-BCRL prevention.^10^ Bioimpedance spectroscopy (BIS) allows for early identification of subclinical extracellular fluid change.^11,12^ Early diagnosis of subclinical BCRL (S-BCRL) coupled with short-course compression therapy has been shown to improve outcomes, suggesting that such early detection may prevent the need for CDP.^13–17^ The primary aim of this study was to determine if subclinical detection of increasing extracellular fluid via BIS and early intervention with four weeks of a compression sleeve and gauntlet resulted in reduced rates of progression to C-BCRL, defined as ≥ 10% arm volume change from baseline requiring CDP, as compared to the same intervention when initiated by increasing arm volume ascertained using circumferential tape measurement (TM).

## Materials and Methods

### Study Design

This multi-center, international, randomized clinical trial (RCT), compared BIS and TM measurements for BCRL surveillance among newly diagnosed breast cancer patients. When patients in either group exceeded pre-established change thresholds from baseline a compression intervention was triggered. The trial was conducted in breast clinics at four sites in Australia and nine sites in the United States. Study procedures were initially approved by the Vanderbilt University Institutional Review Board (IRB) and the Vanderbilt Ingram Center Scientific Review Committee (SRC). Subsequent approvals were obtained by individual site IRBs, and when required institutional SRC’s. Vanderbilt University School of Nursing (VUSN) served as the coordinating/lead site, provided all training, as well as data review, maintenance and analysis. The Research Electronic Data Capture (REDCap) database environment was used for data collection and management.^18^ Fidelity oversite visits and data/safety audits were conducted annually at each of the sites.

### Participants

Presurgical inclusion criteria included women over 18 years old with histologically confirmed newly diagnosed breast cancer (invasive carcinoma or ductal carcinoma in situ (DCIS)) with a planned surgical procedure. Exclusion criteria included: history of breast cancer; neoadjuvant chemotherapy (latter half of recruitment phase of study only as three year follow-up would extend past planned study end); previous radiation to the breast/chest wall/or axilla for any reason; active implanted medical device (e. g., pace maker); medical conditions known to cause swelling; pregnancy; previous arm lymphedema treatment; uncontrolled intercurrent illness; psychiatric illness that would limit compliance; planned bilateral surgery; or an allergy to electrode adhesives or compression fabrics. The study was designed to be reflective of an elevated risk of S-BCRL. Therefore, eligibility was re-evaluated two months (+/-5 days) after surgery prior to randomization, with inclusion criteria of stage I–III invasive breast cancer or DCIS with at least one of the following: mastectomy, axillary treatment (ALND, SLNB with >6 nodes removed), or RNI to the chest wall/breast, axilla, and/or supraclavicular fossa, or taxane-based chemotherapy. Those undergoing bilateral mastectomies were excluded. Written informed consent was obtained from all participants.

### Procedures

Baseline pre-surgical measurements (including both BIS and TM) were completed for all enrolled patients. Patients who remained eligible after the second evaluation were immediately randomized with 1:1 allocation, using a computer-generated, permuted block program (blocks of four) to measurement by either BIS or TM. Centralized randomization, stratified by site, was implemented by VUSN. Due to the difference in appearance of a BIS device or TM, blinding was not possible for the research assistants taking measurements.

Those assigned to the BIS group were assessed using an L-Dex^®^U400 (ImpediMed Limited, Brisbane, Australia). Measurements were conducted by trained research staff, following manufacturer’s instructions and reported in L-Dex^®^ units. Those in the TM group were measured with a Gulick II tape. Using a marked board to facilitate correct tape placement, arms were measured twice at 10 cm increments from the wrist up to 50 cm above the wrist by trained research staff. Arm volume was auto-calculated using a truncated cone formula and the average of the two assessments was used for evaluation.

Initially the intervention trigger for the BIS group was ≥10 L-Dex units in the absence of a >10% volume change from pre-treatment baseline and for the TM group an at-risk arm volume change of ≥5% and <10% compared to baseline and contralateral measurements. In either group a volume change of >10% resulted in direct referral for CDP. In 2016, published studies demonstrated the presence of early-stage C-BCRL with the BIS of seven rather than ten L-Dex units.^19–21^ We verified those findings in a sample of 280 women.^22^ Thus, with IRB and SRC approval the intervention trigger was modified from ≥10 L-Dex units to ≥6.5 L-Dex units for all previously enrolled and subsequent patients assigned to the BIS group.

Postoperative BIS or TM assessments for all endpoints were at 3, 6, 12, 18, 24, and 36 months and following compression intervention. Optional visits at 15 and 21 months were allowed at site PI discretion. Patients that met the intervention trigger for S-BCRL, wore a class 2 (23–32 mmHg, medi flat knit custom or Harmony^®^ circular knit) compression sleeve and gauntlet for four weeks, 12-hours/day. When a participant triggered an intervention, they underwent the alternative measurement, ensuring both BIS and TM measurements were taken for all participants prior to intervention initiation. Regardless of group assignment, if at intervention triggering, intervention completion, or any measurement post-intervention TM volume in the at-risk arm was ≥10% change from baseline (absent a similar change in the non-at-risk arm) the patient was referred for CDP due to BCRL progression and removed from the study.

### Statistical Analysis

We hypothesized that compared to TM detection of subclinical swelling, BIS detection would reduce the rate of lymphedema progression requiring CDP by as much as 20%. Estimates of progression vary widely thus a rate of 50% using TM was used to determine that group sizes of 100 achieved 80% statistical power to detect at least that much difference (2-sided < 0.05). If the observed rates in the TM group were higher or lower than 50%, then the statistical power of a proposed 20% difference would increase and smaller differences could be detected.

Categorical variables were summarized using frequency distributions and compared using Chi-square tests of independence. Median and inter-quartile range were used to summarize continuous study variables; group comparisons were conducted using Mann–Whitney tests. Analyses of progression after triggering used intent-to-treat (ITT) principles (comparison of randomization conditions regardless of whether or not patients completed intervention post-trigger). The primary hypothesis that surveillance and early detection with BIS would result in lower rates of chronic BCRL.

C-BCRL, defined as referral for CDP when there was a ≥ 10% arm volume change from baseline) when using detection via TM was tested using logistic regression. A bootstrapped probability and 95% confidence interval were generated around the parameter estimate using 1000 bootstrap samples. Statistical significance was established as a maximum 2-sided Type I error of 0.05.

## Results

Overall, 1,239 women were recruited between June 24, 2014 and September 11, 2018 with 1,200 enrolled; post-operatively, 963 patients met the inclusion criteria and were randomized (BIS n=482: TM n=481) (see Consort Diagram). The final analysis was performed upon completion of the study (December 31, 2020) on 879 patients (BIS n=442; TM n=437) as 39 (4.2%) patients progressed either at their initial post-randomization visit or between other study visits (BIS n=19; TM n=20, p=.85) and 45 had no valid post-baseline assessments (BIS n=21; TM n=24, p=.64). The sample had a median age of 58.5 years with most being non-Hispanic nor Latina (96.4%) (Table 1). With the exception of patients assigned to TM having a slightly higher rate of smoking (37% vs. 30%, *p* = 0.051), the groups were well balanced for demographic and pathologic features. Clinical characteristics also were well-balanced, with the group of BIS patients having a slightly higher prevalence of Stage I disease than TM (61% vs. 52%). In terms of treatment, the BIS group was comprised of a slightly higher proportion of patients who had SLNB than the TM group (84% vs. 78%, p = 0.011 Table 2). No other statistical or meaningful differences in number of nodes removed, utilization of RNI or taxane-based chemotherapy were observed between groups. Furthermore, within the ultimate sample (those who triggered an intervention) for testing our primary outcome (progression), no significant or meaningful differences between the rates of ALND (30.3% vs. 27.6%, *p* = 0.127) or cancer stage at presentation were observed (*p* = 0.475, Table 3).

**Table 1.** Sociodemographic and Environmental Characteristics at Baseline.

| Characteristic | Overall<br>(N=879) | TM Group<br>(N=437) | BIS Group<br>(N=442) | p |
| --- | --- | --- | --- | --- |
|  | Median [IQR] (N) |  |  |  |
| Age | 58.5 [50, 67] (878) | 58.8 [50, 66] (437) | 58.4 [50, 67] (441) | 0.944 |
| Missing | 1 | 0 | 1 |  |
| Years of Education | 16.0 [12, 16] (873) | 16.0 [12, 16] (435) | 16.0 [12, 16] (438) | 0.444 |
| Missing | 6 | 2 | 4 |  |
|  | n (%) |  |  |  |
| Race |  |  |  | 0.774 |
| Asian | 76 (8.8) | 38 (8.7) | 38 (8.8) |  |
| Black or African American | 67 (7.7) | 31 (7.1) | 36 (8.3) |  |
| White | 678 (78.1) | 345 (79.3) | 333 (76.9) |  |
| Multiracial or Other | 47 (5.4) | 21 (4.8) | 26 (6.0) |  |
| Missing | 11 | 2 | 9 |  |
| Ethnicity |  |  |  | 0.685 |
| Non-Hispanic nor Latina | 805 (96.4) | 399 (96.1) | 406 (96.7) |  |
| Hispanic or Latina | 30 (3.6) | 16 (3.9) | 14 (3.3) |  |
| Missing | 44 | 22 | 22 |  |
| Ever Smoked | 294 (33.5) | 160 (36.6) | 134 (30.4) | 0.051 |
| Ever Drank alcohol | 621 (70.7) | 315 (72.1) | 306 (69.4) | 0.380 |
| Stage of Cancer |  |  |  | 0.058 |
| 0 (DCIS) | 54 (6.1) | 27 (6.2) | 27 (6.1) |  |
| I | 496 (56.4) | 228 (52.2) | 268 (60.6) |  |
| II | 270 (30.7) | 152 (34.8) | 118 (26.7) |  |
| III | 59 (6.7) | 30 (6.9) | 29 (6.6) |  |
| Baseline Assessments |  |  |  |  |
|  | Median [IQR] |  |  |  |
| L-Dex Units | 0.1 [-3.1, 3.2] | 0.1 [-3.2, 3.2] | 0.2 [-3.0, 3.2] | 0.999 |
| Arm Volume |  |  |  |  |
| At-risk Arm (mL) | 1966.4<br>(1694, 2354) | 1968.7<br>[1692, 2342] | 1966.4<br>[1697, 2369] | 0.775 |
| Non-at-risk Arm (mL) | 1970.0<br>[1689, 2371] | 1964.5<br>[1688, 2351] | 1973.9<br>[1689, 2392] | 0.613 |
| Percent Difference | 0.1 [-3.1, 3.2] | 0.2 [-3.2, 3.2] | 0.1 [-2.8, 3.2] | 0.672 |
| BMI | 27.9<br>[24.6, 32.5] | 28.3<br>[24.6, 32.5] | 27.6<br>[24.5, 32.4] | 0.684 |

Note: All participants indicated female gender. (Abbreviations: BIS – Bioimpedance spectroscopy; DCIS –44 Ductal Carcinoma In Situ; BMI – Body Mass Index)

**Table 2:**
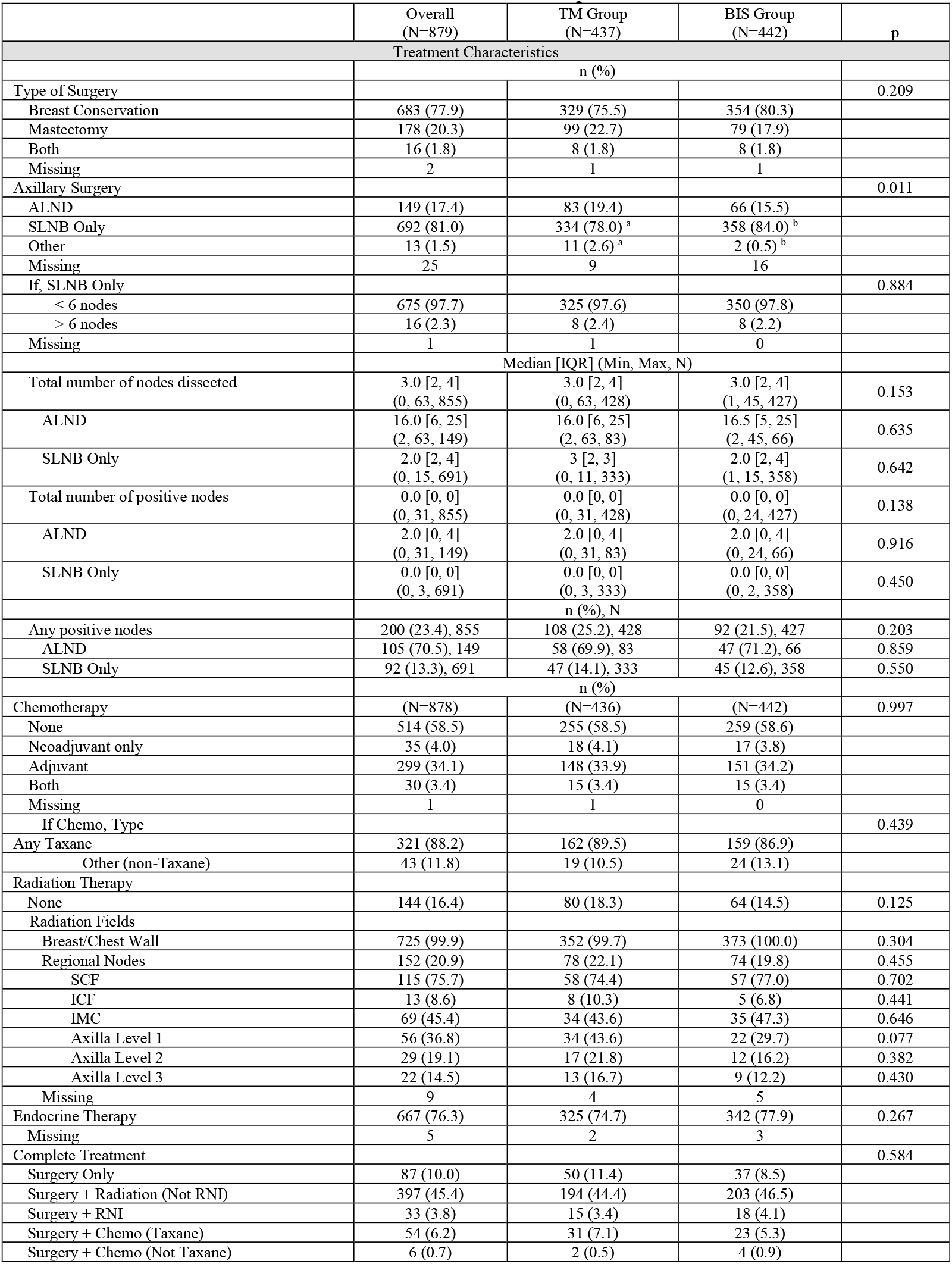
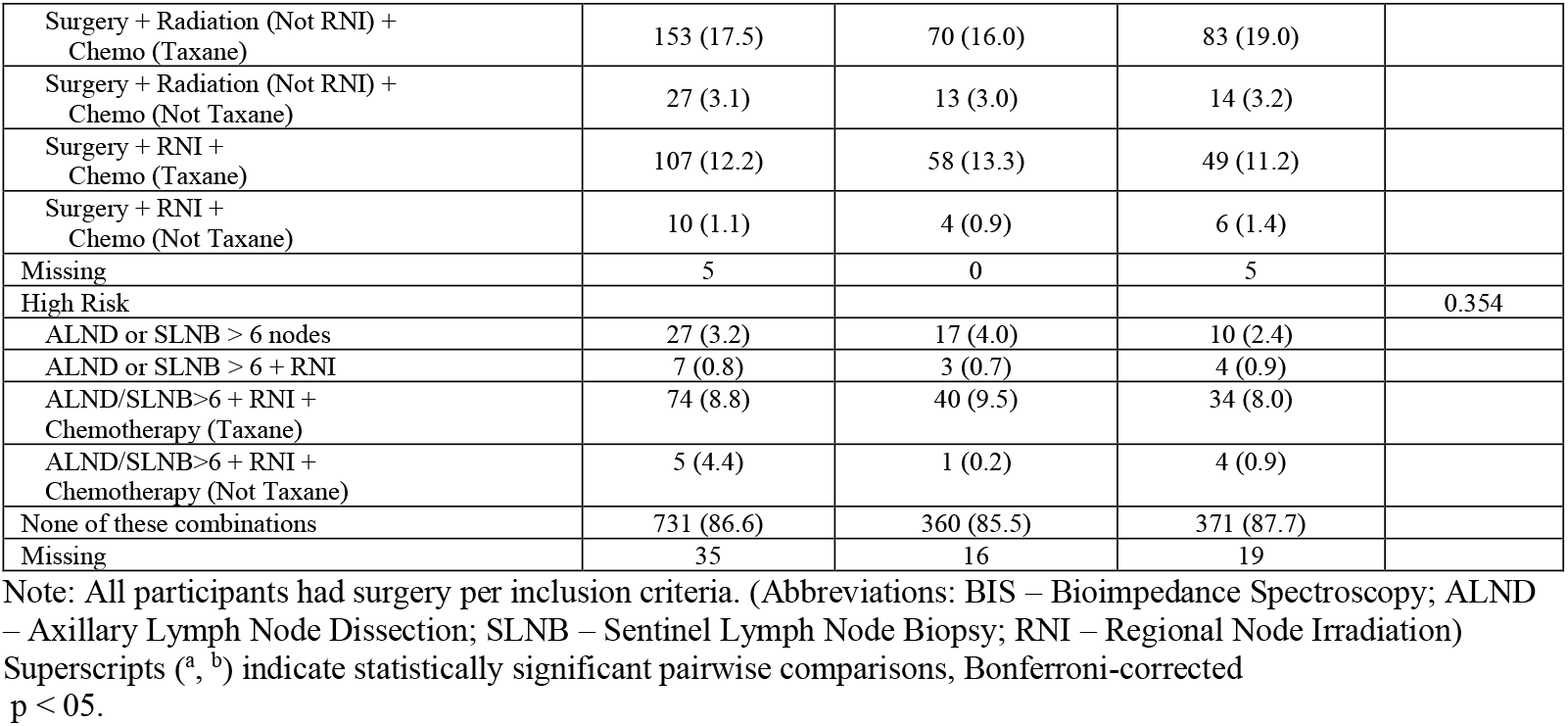
Breast Treatment* Characteristics all Randomized Participants.

|  | Overall<br>(N=879) | TM Group<br>(N=437) | BIS Group<br>(N=442) | p |
| --- | --- | --- | --- | --- |
| **Treatment Characteristics** |  |  |  |  |
|  | n (%) |  |  |  |
| Type of Surgery |  |  |  | 0.209 |
| Breast Conservation | 683 (77.9) | 329 (75.5) | 354 (80.3) |  |
| Mastectomy | 178 (20.3) | 99 (22.7) | 79 (17.9) |  |
| Both | 16 (1.8) | 8 (1.8) | 8 (1.8) |  |
| Missing | 2 | 1 | 1 |  |
| Axillary Surgery |  |  |  | 0.011 |
| ALND | 149 (17.4) | 83 (19.4) | 66 (15.5) |  |
| SLNB Only | 692 (81.0) | 334 (78.0)<sup>a</sup> | 358 (84.0)<sup>b</sup> |  |
| Other | 13 (1.5) | 11 (2.6)<sup>a</sup> | 2 (0.5)<sup>b</sup> |  |
| Missing | 25 | 9 | 16 |  |
| If, SLNB Only |  |  |  | 0.884 |
| ≤ 6 nodes | 675 (97.7) | 325 (97.6) | 350 (97.8) |  |
| > 6 nodes | 16 (2.3) | 8 (2.4) | 8 (2.2) |  |
| Missing | 1 | 1 | 0 |  |
|  | Median [IQR] (Min, Max, N) |  |  |  |
| Total number of nodes dissected | 3.0 [2, 4]<br>(0, 63, 855) | 3.0 [2, 4]<br>(0, 63, 428) | 3.0 [2, 4]<br>(1, 45, 427) | 0.153 |
| ALND | 16.0 [6, 25]<br>(2, 63, 149) | 16.0 [6, 25]<br>(2, 63, 83) | 16.5 [5, 25]<br>(2, 45, 66) | 0.635 |
| SLNB Only | 2.0 [2, 4]<br>(0, 15, 691) | 3 [2, 3]<br>(0, 11, 333) | 2.0 [2, 4]<br>(1, 15, 358) | 0.642 |
| Total number of positive nodes | 0.0 [0, 0]<br>(0, 31, 855) | 0.0 [0, 0]<br>(0, 31, 428) | 0.0 [0, 0]<br>(0, 24, 427) | 0.138 |
| ALND | 2.0 [0, 4]<br>(0, 31, 149) | 2.0 [0, 4]<br>(0, 31, 83) | 2.0 [0, 4]<br>(0, 24, 66) | 0.916 |
| SLNB Only | 0.0 [0, 0]<br>(0, 3, 691) | 0.0 [0, 0]<br>(0, 3, 333) | 0.0 [0, 0]<br>(0, 2, 358) | 0.450 |
|  | n (%), N |  |  |  |
| Any positive nodes | 200 (23.4), 855 | 108 (25.2), 428 | 92 (21.5), 427 | 0.203 |
| ALND | 105 (70.5), 149 | 58 (69.9), 83 | 47 (71.2), 66 | 0.859 |
| SLNB Only | 92 (13.3), 691 | 47 (14.1), 333 | 45 (12.6), 358 | 0.550 |
|  | n (%) |  |  |  |
| Chemotherapy | (N=878) | (N=436) | (N=442) | 0.997 |
| None | 514 (58.5) | 255 (58.5) | 259 (58.6) |  |
| Neoadjuvant only | 35 (4.0) | 18 (4.1) | 17 (3.8) |  |
| Adjuvant | 299 (34.1) | 148 (33.9) | 151 (34.2) |  |
| Both | 30 (3.4) | 15 (3.4) | 15 (3.4) |  |
| Missing | 1 | 1 | 0 |  |
| If Chemo, Type |  |  |  | 0.439 |
| Any Taxane | 321 (88.2) | 162 (89.5) | 159 (86.9) |  |
| Other (non-Taxane) | 43 (11.8) | 19 (10.5) | 24 (13.1) |  |
| Radiation Therapy |  |  |  |  |
| None | 144 (16.4) | 80 (18.3) | 64 (14.5) | 0.125 |
| Radiation Fields |  |  |  |  |
| Breast/Chest Wall | 725 (99.9) | 352 (99.7) | 373 (100.0) | 0.304 |
| Regional Nodes | 152 (20.9) | 78 (22.1) | 74 (19.8) | 0.455 |
| SCF | 115 (75.7) | 58 (74.4) | 57 (77.0) | 0.702 |
| ICF | 13 (8.6) | 8 (10.3) | 5 (6.8) | 0.441 |
| IMC | 69 (45.4) | 34 (43.6) | 35 (47.3) | 0.646 |
| Axilla Level 1 | 56 (36.8) | 34 (43.6) | 22 (29.7) | 0.077 |
| Axilla Level 2 | 29 (19.1) | 17 (21.8) | 12 (16.2) | 0.382 |
| Axilla Level 3 | 22 (14.5) | 13 (16.7) | 9 (12.2) | 0.430 |
| Missing | 9 | 4 | 5 |  |
| Endocrine Therapy | 667 (76.3) | 325 (74.7) | 342 (77.9) | 0.267 |
| Missing | 5 | 2 | 3 |  |
| Complete Treatment |  |  |  | 0.584 |
| Surgery Only | 87 (10.0) | 50 (11.4) | 37 (8.5) |  |
| Surgery + Radiation (Not RNI) | 397 (45.4) | 194 (44.4) | 203 (46.5) |  |
| Surgery + RNI | 33 (3.8) | 15 (3.4) | 18 (4.1) |  |
| Surgery + Chemo (Taxane) | 54 (6.2) | 31 (7.1) | 23 (5.3) |  |
| Surgery + Chemo (Not Taxane) | 6 (0.7) | 2 (0.5) | 4 (0.9) |  |
| Surgery + Radiation (Not RNI) + Chemo (Taxane) | 153 (17.5) | 70 (16.0) | 83 (19.0) |  |
| Surgery + Radiation (Not RNI) + Chemo (Not Taxane) | 27 (3.1) | 13 (3.0) | 14 (3.2) |  |
| Surgery + RNI + Chemo (Taxane) | 107 (12.2) | 58 (13.3) | 49 (11.2) |  |
| Surgery + RNI + Chemo (Not Taxane) | 10 (1.1) | 4 (0.9) | 6 (1.4) |  |
| Missing | 5 | 0 | 5 |  |
| High Risk |  |  |  | 0.354 |
| ALND or SLNB > 6 nodes | 27 (3.2) | 17 (4.0) | 10 (2.4) |  |
| ALND or SLNB > 6 + RNI | 7 (0.8) | 3 (0.7) | 4 (0.9) |  |
| ALND/SLNB>6 + RNI + Chemotherapy (Taxane) | 74 (8.8) | 40 (9.5) | 34 (8.0) |  |
| ALND/SLNB>6 + RNI + Chemotherapy (Not Taxane) | 5 (4.4) | 1 (0.2) | 4 (0.9) |  |
| None of these combinations | 731 (86.6) | 360 (85.5) | 371 (87.7) |  |
| Missing | 35 | 16 | 19 |  |

Note: All participants had surgery per inclusion criteria. (Abbreviations: BIS – Bioimpedance Spectroscopy; ALND – Axillary Lymph Node Dissection; SLNB – Sentinel Lymph Node Biopsy; RNI – Regional Node Irradiation) Superscripts (<sup>a</sup>, <sup>b</sup>) indicate statistically significant pairwise comparisons, Bonferroni-corrected $p < 0.05$ .

**Table 3:**
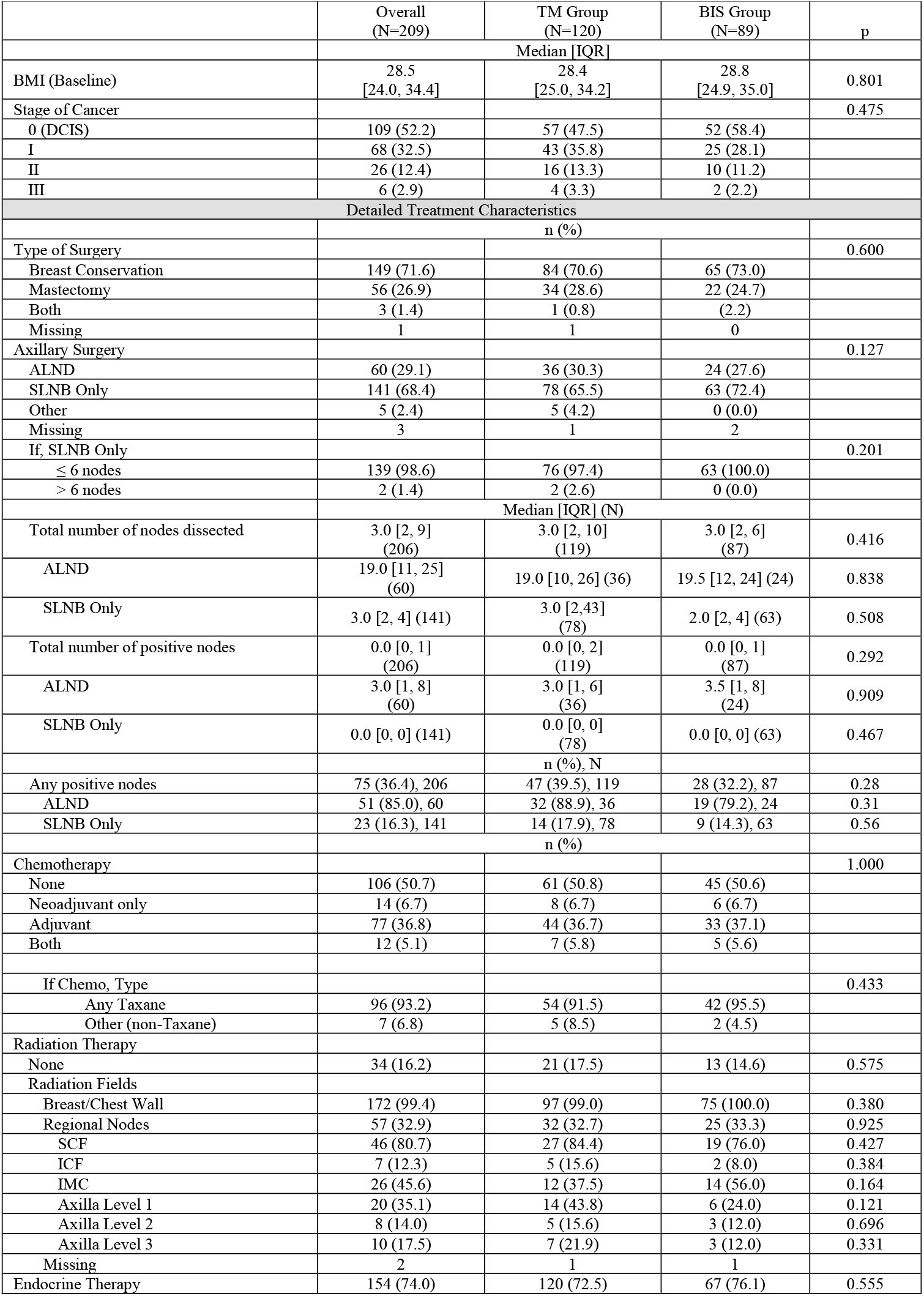
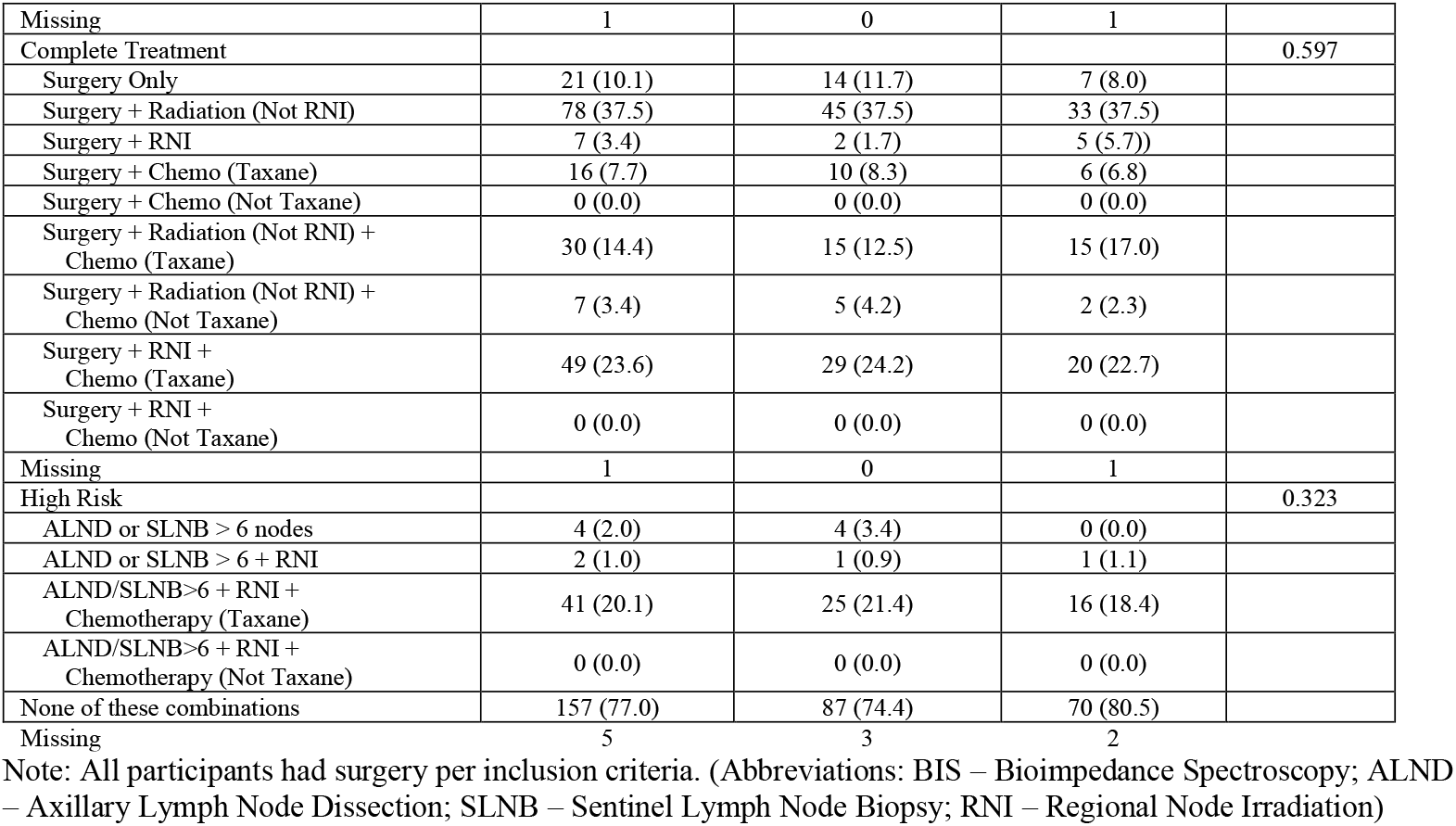
Critical Clinical and Breast Treatment Characteristics of Sample Triggering Intervention (N=209)

|  | Overall<br>(N=209) | TM Group<br>(N=120) | BIS Group<br>(N=89) | p |
| --- | --- | --- | --- | --- |
|  | Median [IQR] |  |  |  |
| BMI (Baseline) | 28.5<br>[24.0, 34.4] | 28.4<br>[25.0, 34.2] | 28.8<br>[24.9, 35.0] | 0.801 |
| Stage of Cancer |  |  |  | 0.475 |
| 0 (DCIS) | 109 (52.2) | 57 (47.5) | 52 (58.4) |  |
| I | 68 (32.5) | 43 (35.8) | 25 (28.1) |  |
| II | 26 (12.4) | 16 (13.3) | 10 (11.2) |  |
| III | 6 (2.9) | 4 (3.3) | 2 (2.2) |  |
| Detailed Treatment Characteristics |  |  |  |  |
|  | n (%) |  |  |  |
| Type of Surgery |  |  |  | 0.600 |
| Breast Conservation | 149 (71.6) | 84 (70.6) | 65 (73.0) |  |
| Mastectomy | 56 (26.9) | 34 (28.6) | 22 (24.7) |  |
| Both | 3 (1.4) | 1 (0.8) | (2.2) |  |
| Missing | 1 | 1 | 0 |  |
| Axillary Surgery |  |  |  | 0.127 |
| ALND | 60 (29.1) | 36 (30.3) | 24 (27.6) |  |
| SLNB Only | 141 (68.4) | 78 (65.5) | 63 (72.4) |  |
| Other | 5 (2.4) | 5 (4.2) | 0 (0.0) |  |
| Missing | 3 | 1 | 2 |  |
| If, SLNB Only |  |  |  | 0.201 |
| ≤ 6 nodes | 139 (98.6) | 76 (97.4) | 63 (100.0) |  |
| > 6 nodes | 2 (1.4) | 2 (2.6) | 0 (0.0) |  |
|  | Median [IQR] (N) |  |  |  |
| Total number of nodes dissected | 3.0 [2, 9]<br>(206) | 3.0 [2, 10]<br>(119) | 3.0 [2, 6]<br>(87) | 0.416 |
| ALND | 19.0 [11, 25]<br>(60) | 19.0 [10, 26] (36) | 19.5 [12, 24] (24) | 0.838 |
| SLNB Only | 3.0 [2, 4] (141) | 3.0 [2, 43]<br>(78) | 2.0 [2, 4] (63) | 0.508 |
| Total number of positive nodes | 0.0 [0, 1]<br>(206) | 0.0 [0, 2]<br>(119) | 0.0 [0, 1]<br>(87) | 0.292 |
| ALND | 3.0 [1, 8]<br>(60) | 3.0 [1, 6]<br>(36) | 3.5 [1, 8]<br>(24) | 0.909 |
| SLNB Only | 0.0 [0, 0] (141) | 0.0 [0, 0]<br>(78) | 0.0 [0, 0] (63) | 0.467 |
|  | n (%), N |  |  |  |
| Any positive nodes | 75 (36.4), 206 | 47 (39.5), 119 | 28 (32.2), 87 | 0.28 |
| ALND | 51 (85.0), 60 | 32 (88.9), 36 | 19 (79.2), 24 | 0.31 |
| SLNB Only | 23 (16.3), 141 | 14 (17.9), 78 | 9 (14.3), 63 | 0.56 |
|  | n (%) |  |  |  |
| Chemotherapy |  |  |  | 1.000 |
| None | 106 (50.7) | 61 (50.8) | 45 (50.6) |  |
| Neoadjuvant only | 14 (6.7) | 8 (6.7) | 6 (6.7) |  |
| Adjuvant | 77 (36.8) | 44 (36.7) | 33 (37.1) |  |
| Both | 12 (5.1) | 7 (5.8) | 5 (5.6) |  |
| If Chemo, Type |  |  |  | 0.433 |
| Any Taxane | 96 (93.2) | 54 (91.5) | 42 (95.5) |  |
| Other (non-Taxane) | 7 (6.8) | 5 (8.5) | 2 (4.5) |  |
| Radiation Therapy |  |  |  |  |
| None | 34 (16.2) | 21 (17.5) | 13 (14.6) | 0.575 |
| Radiation Fields |  |  |  |  |
| Breast/Chest Wall | 172 (99.4) | 97 (99.0) | 75 (100.0) | 0.380 |
| Regional Nodes | 57 (32.9) | 32 (32.7) | 25 (33.3) | 0.925 |
| SCF | 46 (80.7) | 27 (84.4) | 19 (76.0) | 0.427 |
| ICF | 7 (12.3) | 5 (15.6) | 2 (8.0) | 0.384 |
| IMC | 26 (45.6) | 12 (37.5) | 14 (56.0) | 0.164 |
| Axilla Level 1 | 20 (35.1) | 14 (43.8) | 6 (24.0) | 0.121 |
| Axilla Level 2 | 8 (14.0) | 5 (15.6) | 3 (12.0) | 0.696 |
| Axilla Level 3 | 10 (17.5) | 7 (21.9) | 3 (12.0) | 0.331 |
| Missing | 2 | 1 | 1 |  |
| Endocrine Therapy | 154 (74.0) | 120 (72.5) | 67 (76.1) | 0.555 |
| Missing | 1 | 0 | 1 |  |
| Complete Treatment |  |  |  | 0.597 |
| Surgery Only | 21 (10.1) | 14 (11.7) | 7 (8.0) |  |
| Surgery + Radiation (Not RNI) | 78 (37.5) | 45 (37.5) | 33 (37.5) |  |
| Surgery + RNI | 7 (3.4) | 2 (1.7) | 5 (5.7) |  |
| Surgery + Chemo (Taxane) | 16 (7.7) | 10 (8.3) | 6 (6.8) |  |
| Surgery + Chemo (Not Taxane) | 0 (0.0) | 0 (0.0) | 0 (0.0) |  |
| Surgery + Radiation (Not RNI) + Chemo (Taxane) | 30 (14.4) | 15 (12.5) | 15 (17.0) |  |
| Surgery + Radiation (Not RNI) + Chemo (Not Taxane) | 7 (3.4) | 5 (4.2) | 2 (2.3) |  |
| Surgery + RNI + Chemo (Taxane) | 49 (23.6) | 29 (24.2) | 20 (22.7) |  |
| Surgery + RNI + Chemo (Not Taxane) | 0 (0.0) | 0 (0.0) | 0 (0.0) |  |
| Missing | 1 | 0 | 1 |  |
| High Risk |  |  |  | 0.323 |
| ALND or SLNB > 6 nodes | 4 (2.0) | 4 (3.4) | 0 (0.0) |  |
| ALND or SLNB > 6 + RNI | 2 (1.0) | 1 (0.9) | 1 (1.1) |  |
| ALND/SLNB>6 + RNI + Chemotherapy (Taxane) | 41 (20.1) | 25 (21.4) | 16 (18.4) |  |
| ALND/SLNB>6 + RNI + Chemotherapy (Not Taxane) | 0 (0.0) | 0 (0.0) | 0 (0.0) |  |
| None of these combinations | 157 (77.0) | 87 (74.4) | 70 (80.5) |  |
| Missing | 5 | 3 | 2 |  |

Note: All participants had surgery per inclusion criteria. (Abbreviations: BIS – Bioimpedance Spectroscopy; ALND – Axillary Lymph Node Dissection; SLNB – Sentinel Lymph Node Biopsy; RNI – Regional Node Irradiation)

Median follow-up was 32.9 months (IQR = 22, 35); with a trend for longer follow-up in the BIS arm than TM (33.1 months; IQR=25, 35 vs. 32.7 months; IQR=20, 35, *p* = 0.054). A lower proportion of BIS patients triggered an intervention (BIS 20.1%, n=89 of 442; 95% CI, 16.3%, 24.0% vs. TM 27.5%, n=120 of 437; 95% CI, 22.9%, 31.7%; *p* = 0.011) and median months from randomization to intervention triggering was longer in the BIS group than the TM group (9.7 months; 95% CI, 8.2-12.6 vs. 3.9 months; 95% CI, 2.8-4.5, *p* = 0.001, Table 4). Of the 209 patients, BIS (n=89) and TM (n=120) who triggered an intervention, 30 (14.4%) progressed after intervention. Of those patients, no difference between the groups was observed in intervention completion rates (BIS 92.1%, n=7 vs. TM 95.0%, n=6; *p* = 0.396).

**Table 4:** Summary of Trigger and Progression by Surveillance Group.

|  | All Patients | TM Group | BIS Group | p |
| --- | --- | --- | --- | --- |
|  | N=918 <sup>a</sup> | N=457 | N=461 |  |
|  | n (%) | n (%) | n (%) |  |
| Progressed Before Intervention | 39 (4.2) | 20 (4.4) | 19 (4.1) | 0.848 |
| Sample for Potential Trigger | N=879 | N=437 | N=442 |  |
|  | Median (IQR) | Median (IQR) | Median (IQR) |  |
| Follow-up (months) <sup>b</sup> | 32.9 (22.7, 34.3) | 32.7 (20.8, 34.3) | 33.1 (25.8,34.7) | 0.054 |
|  | n (%) | n (%) | n (%) |  |
| Triggered | 209 (23.8) | 120 (27.5) | 89 (20.1) | 0.011 |
|  | Median (IQR) | Median (IQR) | Median (IQR) |  |
| Time from Randomization to Trigger (months) | 5.1 (1.3, 15.6) | 3.9 (1.0, 11.6) | 9.7 (3.6, 18.2) | 0.001 |
|  | N=209 | N=120 | N=89 |  |
|  | n (%) | n (%) | n (%) |  |
| Progression <sup>c</sup> | 30 (14.4) | 23 (19.2) | 7 (7.9) | 0.016 <sup>d</sup> |
|  | Median (Min, Max) | Median (Min, Max) | Median (Min, Max) |  |
| Time from Trigger to Progression (months) | 9.6 (0.7, 31.9) | 10.7 (1.4, 31.9) | 4.9 (0.7, 15.2) | 0.100 |

(Abbreviations: BIS – Bioimpedance Spectroscopy)
<sup>a</sup> Sample with any data post-treatment
<sup>b</sup> Months from randomization to last assessment using either tape or BIS
<sup>c</sup> Progression to criteria for being off-study without the interim intervention (subclinical) threshold
<sup>d</sup> Reduction 11.3%; 95% CI, 2.3% to 20.3%; OR = 0.36; 95% CI, 0.11 to 0.77, $p = 0.016$ , RR = 0.41; 95% CI, 0.13 to 0.81).

Following intervention, patients in the BIS group were less likely to progress to CDP than those in the TM group (7.9%, n=7 of 89 vs. 19.2%, n=23 of 120; absolute reduction 11.3%; 95% CI, 2.3%-20.3%; OR=0.36, 95% CI=0.11–0.77, *p* = 0.016). Relative rates of progression were reduced by approximately 59% in the BIS arm (RR=0.41; 95% CI, 0.13-0.81). Median follow-up months between trigger and progression for the TM or BIS groups were not statistically significantly different (BIS: 4.9 months; 95% CI, 0.7-13.9 vs. TM: 10.7 months; 95% CI, 4.9 to 17.0, *p* = 0.100, Table 4). An analysis of progression that included only the patients who completed the intervention resulted in similar findings (BIS 8.5%, n=7 of 82 vs. TM: 20.2%, n=23 of 114, absolute reduction 11.7%; 95% CI, 2.1%-21.3%, *p* = 0.018). While the randomized groups who triggered were well-balanced in terms of known risk factors for progression to CDP with no statistically significant differences between them, they certainly were not “case/control matched” or “equivalent’. To account for the possibility that one of those factors rather than the triggering approach accounted for the difference in progression rates, risk-adjusted analyses were conducted. As shown in Table 5, while some of those increased the likelihood of progression to CDP (cancer stage, *p* = 0.002; ALND, *p* < 0.001; any positive nodes removed, *p* = 0.007; chemotherapy, *p* = 0.005; a combination of high-risk treatments, *p* = 0.031), after adjusting for those factors the estimates of the effects of using BIS as the BCRL detection method remained essentially equivalent for all adjustments (bootstrapped ORs of 0.35 to 0.37, with *p* values remaining between 0.01 and 0.03 for effect of BIS, Table 5).

**Table 5:** Risk Adjusted Progression.

|  | **OR** | **95% C.I.** | **p** |
| --- | --- | --- | --- |
| BIS (BMI, p = 0.288) | 0.36 | 0.14 - 0.88 | 0.017 |
| BIS (Cancer stage, p = 0.002) | 0.35 | 0.13 - 0.90 | 0.022 |
| BIS (Mastectomy, p = 0.298) | 0.36 | 0.14 - 0.88 | 0.014 |
| BIS (ALND, p < 0.001) | 0.36 | 0.14 – 0.92 | 0.021 |
| BIS (# nodes removed, p = 0.002) | 0.35 | 0.14 – 0.90 | 0.029 |
| BIS (# positive nodes, p = 0.061) | 0.36 | 0.14 – 0.89 | 0.023 |
| BIS (Any positive nodes, p = 0.009) | 0.39 | 0.15 – 0.96 | 0.022 |
| BIS (Any chemo, p = 0.002) | 0.35 | 0.13 – 0.86 | 0.018 |
| BIS (If chemo, Taxane, N=103, p = 0.276) | 0.31 | 0.10 – 0.92 | 0.024 |
| BIS (Any radiation, p = 0.330) | 0.36 | 0.14 – 0.90 | 0.019 |
| BIS (If radiation, RNI, N=171, p = 0.098) | 0.23 | 0.07 – 0.71 | 0.009 |
| BIS (High Risk Tx Combination, p = 0.031) | 0.37 | 0.15 – 0.93 | 0.023 |

Bootstrapped bias-corrected parameter estimates (OR – odds ratio), confidence intervals, and *p* values for effect of BIS (compared to TM) on the likelihood of progression to CDP after adjusting for the effect of each of the specified known risks for progression. Each of the risks and the probability of their effect on progression that was controlled for in each analyses are shown in ( ).

Adverse event information was available for the 963 randomized patents. The overall number of study-related adverse events was <1% (n=3). Two reports were of Grade 1 skin itching/tingling/redness during the compression intervention. One patient reported perceived swelling of upper arm during the intervention, but physical examination and measurement did not confirm physiological swelling. There were no study related serious adverse events.

## Discussion

The results from this large, multi-center, RCT trial add to the prior reported data on this topic supporting that use of BIS for prospective BCRL screening coupled with early intervention of a short four-week well-fitted compression sleeve and gauntlet reduces progression to C-BCRL as compared to TM. ^23,24^ Specifically, use of BIS as part of prospective BCRL surveillance, coupled with early compression sleeve and gauntlet intervention, significantly reduced C-BCRL (progression to CDP), (7.9% vs 19.2%, P=.016) as compared to TM. Analyses including patients who did not complete intervention after triggering, as well as just those who completed intervention both supported this significance. This represents a clinically significant outcome, offering clinicians the ability to use this approach to reduce a patient’s risk of developing C-BCRL and potentially affords payers the option of a low-cost, conservative compression sleeve intervention instead of expensive treatment such as physical therapy, bypass, and CDP.

This RCT provides level I evidence as compared to previous studies with limited numbers of patients that did not use S-BCRL definitions.^12,25^ This is supported by a meta-analysis of more than 67,000 women and 50 studies that found that use of BIS reduced annualized and cumulative incidence of C-BCRL as compared to TM or background studies, though not controlling for intervention protocols as the current study did.^26^ These consistent significant findings are likely related to the ability of BIS to detect an increase in extracellular fluid, as opposed to TM’s ability to only detect an increase in whole arm volume. Thus, BIS serves as a better screening method to determine who will best benefit from a prevention intervention and achieve reversal of the S-BCRL process that can lead to C-BCRL.^23,24^ BIS is more specific for lymphedema detection than TM as it had fewer triggers and longer times to intervention trigger. The lower rates of C-BCRL in the BIS group would therefore be secondary to more accurate detection and identification of patients with S-BCRL, rather than earlier intervention per se as the time to trigger is significantly longer.

A critically important issue regarding the robustness of our findings is that not only did we conduct a rigorous intent-to-treat analysis of the randomized groups, we also risk-adjusted for those factors known to increase the likelihood of progression. Randomization insured patients had equal opportunity for both types of assessment however “equivalence” of any other factor is not assured. Both groups were well-balanced in terms of high-risk factors for progression (no statistically significant differences). Our risk-adjustment analysis confirmed that most of those factors (cancer stage, total number of nodes removed, any positive nodes removed, chemotherapy, and a combination of high-risk treatments) did statistically significantly increase the likelihood of progression. The parameter estimates for the reduction in likelihood of progression with BIS surveillance remained stable (ORs of 0.35-0.37, P=0.01-0.03, Table 5), confirming the balanced nature of the groups. Thus, the statistically and clinically significant positive outcome experienced by patients in the BIS group was related to the screening method and its detection of changes in extracellular fluid.

BIS is clearly a valuable tool for patient management. Our findings are consistent with previous prospective studies which found that use of BIS as part of prospective surveillance resulted in a persistent C-BCRL rate of only 6%.^26^ The final results of the study, confirm the significantly lower rates of trigger with BIS as compared to TM (20.7% vs 27.5%, *P*=.011) as seen at the interim analysis. ^28, 29, 30^ Critical knowledge to inform the optimal method of prospective surveillance was also generated in this study. Intensive training, a standardized measurement protocol, and annual fidelity oversight for both measurements were undertaken. While the BIS protocol can be easily replicated in clinical settings, the rigor of the TM protocol for this study exceeded what is practical in most clinics. Thus, BIS may offer even more benefit across clinical settings than what was demonstrated in this study.

The trial was amended during enrollment to reflect a lower L-Dex trigger.^21^ Despite this change, TM still generated more intervention triggers. Though intra and inter-observer variability with TM may have contributed to the greater number of triggers in the TM group; the required training and annual fidelity visits were intended to limit this risk as much as possible.

With respect to time to trigger, a difference remained between the two techniques though the difference was smaller than in the interim analysis. This raises questions as to whether TM is capturing post-surgical/radiation inflammatory soft tissue changes that BIS does not capture, rather than lymphedema. If true, then patient’s may be diagnosed with lymphedema, when they do not have it and experience unnecessary psychological distress.

Although this study has many strengths, some limitations do exist. While high-risk features and cancer treatment techniques were well balanced between arms and the statistical analyses controlled for those known and/or potential confounding effects, there may exist effects of those features and/or treatments that could not be accounted for in this trial. Furthermore, this RCT compared BIS to TM and as such, extrapolation to other BCRL diagnostics is not possible. Additionally, there was not a no intervention control arm; any such an arm would eliminate equipoise in the study, was unlikely to be approved by IRBs given the data available regarding C-BCRL prevention, and there was potential for negative impact upon recruitment and retention, given the level of subject burden for a 3-year study commitment should one group not be offered the intervention. The ALND rate for the study was 17%; this is consistent with modern practice as many patients who previously received ALND will now receive RNI instead.^30^ However, strengths of this study include the design, absence of clinically relevant differences in sociodemographic characteristics between groups prior to treatment, and length of follow-up.

## Conclusion

Prospective surveillance conducted over 3-years post operatively that identified and treated S-BCRL improved patient outcomes. Given the large scale, long-term follow-up and randomized nature of the trial, these statistically significant results demonstrate that BIS screening should be a standard approach for prospective BCRL surveillance. BIS as compared to TM, provides a more precise identification of patients likely to benefit from an early compression intervention.

## Supporting information

equator checklist

## Data Availability

The study protocol will be made available at https://nursing.vanderbilt.edu/research/projects/pdf/preventstudy.pdf. The deidentified individual participant data that support the findings will be available at ftp.impedimed.com following an embargo from the date of publication to 12-31-2022 to allow for commercialization of research findings. Data will not be available after 12-31-2023. The ftp site can be accessed using an ftp client such as FileZilla. Username and password can be obtained by contacting

https://ftp.impedimed.com

## Acknowledgements

The authors thank the participants in the study for giving of their time for 3 years to the study.

## Authors Contributions

*Dietrich, Ridner, and Shah:* Conceptualization and writing original draft:

*All authors contributed equally:* Methodology, resources, writing-review-editing.

*Dietrich and Ridner:* Funding Acquisition, authors verifying underlying data, data curation, formal analysis.

*Dietrich:* Visualization.

*Dietrich, Ridner, and Shah:* Validation.

*Dietrich:* Software.

*Abramson, Boyages, Elder, French, Hughes, Hsu, Koelmeyer, Moore, Ngui and Ridner:*

Investigation and project administration.

---

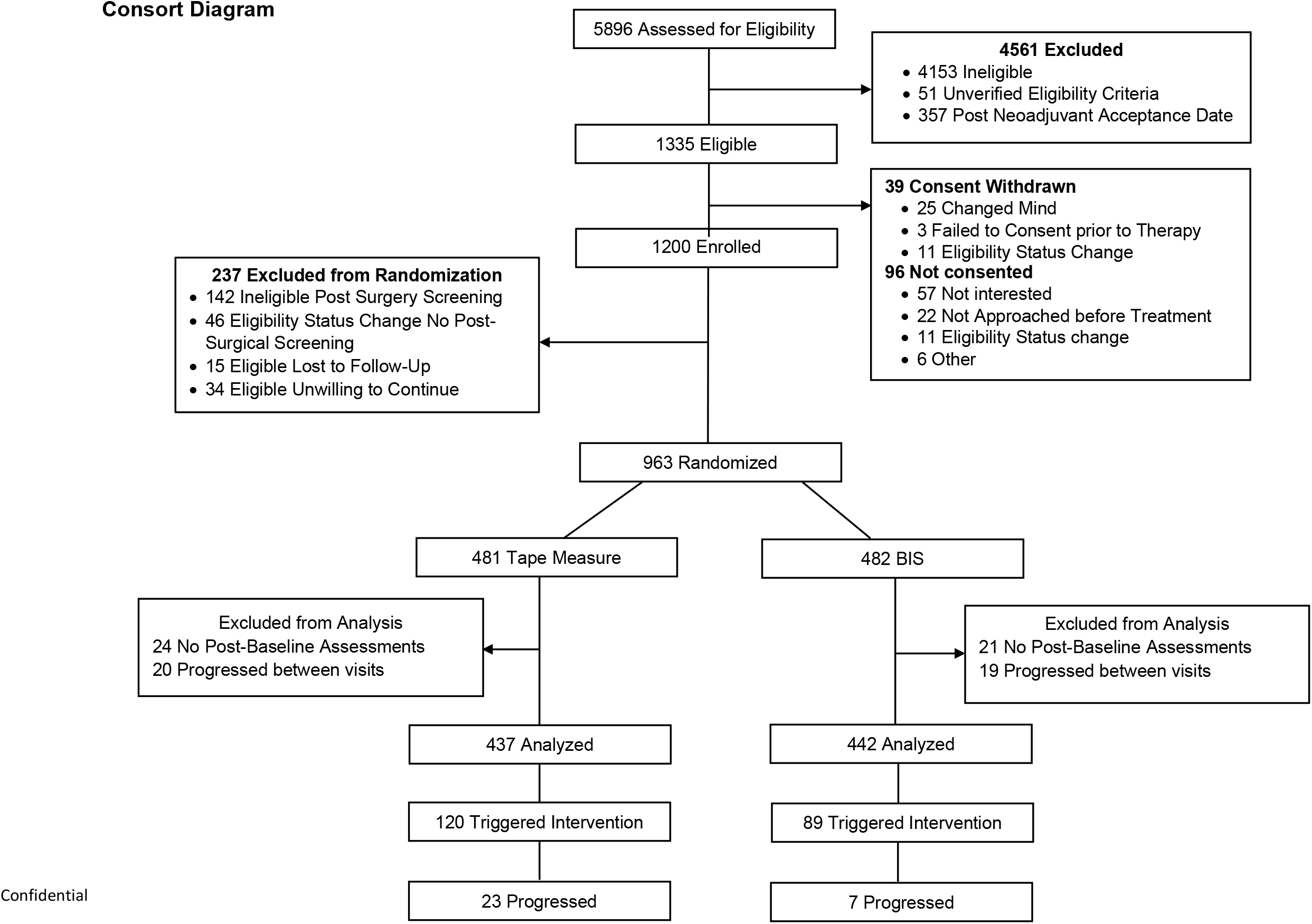

## References

1. Breast Cancer Research Foundation. Breast Cancer Statistics and Resources. 2020. https://www.bcrf.org/breast-cancer-statistics-and-resources (accessed Accessed October 22, 2020).

2. Ashikaga T, Krag DN, Land SR, et al. Morbidity results from the NSABP B-32 trial comparing sentinel lymph node dissection versus axillary dissection. J Surg Oncol 2010; 102(2): 111–8.

3. Coen JJ, Taghian AG, Kachnic LA, Assaad SI, Powell SN. Risk of lymphedema after regional nodal irradiation with breast conservation therapy. Int J Radiat Oncol Biol Phys 2003; 55(5): 1209–15.

4. Vignes Sp, Porcher Rl, Arrault M, Dupuy A. Long-term management of breast cancerrelated lymphedema after intensive decongestive physiotherapy. Breast Cancer Research And Treatment 2007; 101(3): 285–90.

5. Gartner R, Jensen MB, Kronborg L, Ewertz M, Kehlet H, Kroman N. Self-reported arm-lymphedema and functional impairment after breast cancer treatment—a nationwide study of prevalence and associated factors. Breast 2010; 19.

6. Sezgin Ozcan D, Dalyan M, Unsal Delialioglu S, Duzlu U, Polat CS, Koseoglu BF. Complex Decongestive Therapy Enhances Upper Limb Functions in Patients with Breast Cancer-Related Lymphedema. Lymphat Res Biol 2018; 16(5): 446–52.

7. Deutsch M, Land S, Begovic M, Sharif S. The incidence of arm edema in women with breast cancer randomised on the national surgical adjuvant breast and bowel project study B-04 to radical mastectomy versus total mastectomy and radiotherapy versus total mastectomy alone. Int J Radiation Oncology Biol Phys 2008; 70(4): 1020–4.

8. Chen YW, Tsai HJ, Hung HC, Tsauo JY. Reliability study of measurements for lymphedema in breast cancer patients. Am J Phys Med Rehabil 2008; 87(1): 33–8.

9. McLaughlin S, Wright M, Morris K, et al. Prevalence of lymphedema in women with breast cancer 5 years after sentinal lymph node biopsy or axillary dissection: objective measurements. Journal of Clinical Oncology 2008; 26(32): 5213–9.

10. Cornish BH, Chapman M, Hirst C, et al. Early diagnosis of lymphedema using multiple frequency bioimpedance. Lymphology 2001; 34(1): 2–11.

11. Shah C, Arthur DW, Wazer D, Khan A, Ridner S, Vicini F. The impact of early detection and intervention of breast cancer-related lymphedema: a systematic review. Cancer Med 2016.

12. Laidley M, Alison, Anglin M, Beth. The Impact of L-Dex® Measurements in Assessing Breast Cancer Related Lymphedema (BCRL) as Part of Routine Clinical Practice. Frontiers in oncology 2016; 6(192).

13. Stout Gergich NL, Pfalzer LA, McGarvey C, Springer B, Gerber LH, Soballe P. Preoperative assessment enables the early diagnosis and successful treatment of lymphedema. Cancer 2008; 112(12): 2809–19.

14. Box RC, Reul-Hirche HM, Bullock-Saxton JE, Furnival CM. Physiotherapy after breast cancer surgery: results of a randomised controlled study to minimise lymphoedema. Breast Cancer Res Treat 2002; 75(1): 51–64.

15. Torres Lacomba M, Yuste Sanchez MJ, Zapico Goni A, et al. Effectiveness of early physiotherapy to prevent lymphoedema after surgery for breast cancer: randomised, single blinded, clinical trial. BMJ 2010; 340: b5396.

16. Dayes IS, Whelan TJ, Julian JA, et al. Randomized trial of decongestive lymphatic therapy for the treatment of lymphedema in women with breast cancer. J Clin Oncol 2013; 31(30): 3758–63.

17. Soran A, Ozmen T, McGuire KP, et al. The importance of detection of subclinical lymphedema for the prevention of breast cancer-related clinical lymphedema after axillary lymph node dissection: A prospective observational study. Lymphat Res Biol 2014; 12(4): 289–94.

18. Harris PA, Taylor R, Thielke R, Payne J, Gonzalez N, Conde JG. Research electronic data capture (REDCap)--a metadata-driven methodology and workflow process for providing translational research informatics support. Journal of biomedical informatics 2009; 42(2): 377–81.

19. Lahtinen T, Seppala J, Viren T, Johansson K. Experimental and Analytical Comparisons of Tissue Dielectric Constant (TDC) and Bioimpedance Spectroscopy (BIS) in Assessment of Early Arm Lymphedema in Breast Cancer Patients after Axillary Surgery and Radiotherapy. Lymphat Res Biol 2015; 13(3): 176–85.

20. Fu MR, Cleland CM, Guth AA, et al. L-dex ratio in detecting breast cancer-related lymphedema: reliability, sensitivity, and specificity. Lymphology 2013; 46(2): 85–96.

21. Dylke ES, Schembri GP, Bailey DL, et al. Diagnosis of upper limb lymphedema: development of an evidence-based approach. Acta Oncol 2016: 1–7.

22. Ridner SH, Dietrich MS, Spotanski K, et al. A Prospective Study of L-Dex Values in Breast Cancer Patients Pretreatment and Through 12 Months Postoperatively. Lymphat Res Biol 2018; 16(5): 435–41.

23. Whitworth PW, Shah C, Vicini F, Cooper A. Preventing Breast Cancer-Related Lymphedema in High-Risk Patients: The Impact of a Structured Surveillance Protocol Using Bioimpedance Spectroscopy. Front Oncol 2018; 8: 197.

24. Kaufman DI, Shah C, Vicini FA, Rizzi M. Utilization of bioimpedance spectroscopy in the prevention of chronic breast cancer-related lymphedema. Breast Cancer Res Treat 2017; 166(3): 809–15.

25. Barrio AV, Eaton A, Frazier TG. A Prospective Validation Study of Bioimpedance with Volume Displacement in Early-Stage Breast Cancer Patients at Risk for Lymphedema. Ann Surg Oncol 2015; 22 Suppl 3(0 3): S370–5.

26. Shah C, Zambelli-Weiner A, Delgado N, Sier A, Bauserman R, Nelms J. The impact of monitoring techniques on progression to chronic breast cancer-related lymphedema: a metaanalysis comparing bioimpedance spectroscopy versus circumferential measurements. Breast Cancer Res Treat 2020; 185(3): 709–40.

27. Kilgore LJ, Korentager SS, Hangge AN, et al. Reducing Breast Cancer-Related Lymphedema (BCRL) Through Prospective Surveillance Monitoring Using Bioimpedance Spectroscopy (BIS) and Patient Directed Self-Interventions. Ann Surg Oncol 2018; 25(10): 2948–52.

28. Ridner SH, Dietrich MS, Cowher MS, et al. A Randomized Trial Evaluating Bioimpedance Spectroscopy Versus Tape Measurement for the Prevention of Lymphedema Following Treatment for Breast Cancer: Interim Analysis. Ann Surg Oncol 2019; 26(10): 3250–9.

29. Ridner SH, Shah C, Boyages J, et al. L-Dex, arm volume, and symptom trajectories 24 months after breast cancer surgery. Cancer Med 2020; 9(14): 5164–73.

30. Donker M, van Tienhoven G, Straver ME, et al. Radiotherapy or surgery of the axilla after a positive sentinel node in breast cancer (EORTC 10981-22023 AMAROS): a randomised, multicentre, open-label, phase 3 non-inferiority trial. Lancet Oncol 2014; 15(12): 1303–10.

